# The Changing Global Epidemiology of Re-emerging Human Monkeypox Virus Infection: A Systematic Review

**DOI:** 10.1101/2022.12.09.22283261

**Authors:** Sunder Sham, FNU Sapna, FNU Anjali, Sanjay Kumar, Vivek Podder, Soumya Jaladi, Ahmed Bendari, Reham Al-Refai, Manal Mohammad Baloch, Mohammed Abdelwahed, FNU Kiran, Hansini Laharwani

## Abstract

**Background:** Human monkeypox (MPXV) virus infection, an emerging zoonotic disease caused by an orthopoxvirus, leads to smallpox-like disease. Human MPXV infection was first reported in 1970 in the Democratic Republic of Congo (DRC). Before April 2022, MPXV cases were endemic and seldom reported outside African regions; but recent global outbreaks of MPXV are concerning. We aimed to investigate the epidemiology of emerging human monkeypox virus infection including the number of suspected, confirmed, and fatal cases as well as risk factors for contracting monkeypox infection.

**Design:** We performed a systematic review of peer-reviewed literature by following Preferred Reporting Items for Systematic Reviews and Meta-Analyses (PRISMA) guideline. An electronic database search (PubMed, online Willey library, science direct) was undertaken. For monkeypox related studies, we included 25 peer-reviewed articles from 2018 and 2022 and data were extracted to inform current evidence on the cases and public health risk factors for developing infection, and public health advice.

**Results:** Our reports show a rapid rise of MPVX cases in highly endemic African regions after the 1970s, spread to other countries, and increased median age from young children to young adults. Cessation of smallpox vaccination might be one of the factors responsible for the findings. Till 2022 genomic sequences of ten MPXV strains, associated with the recent countrywide outbreak, have been determined. While West African Clade is mostly implicated in the recent viral surge, data were insufficient to determine which mutation contributed to increased transmissibility. In DRC, sleeping on the floor was significantly (odds ratio [OR] 6.1, 95% confidence interval [CI] 1.2-31.6) associated with contracting MPVX *while eating or processing animal foods was not a significant risk factor*. In the United States, cleaning cages, bedding sick animals (OR 5.3, 95% CI 1.4-20.7), or touching infected animals (OR 4.0, 95% CI 1.2-13.4), daily sick animal exposure (OR 4.0, 95% CI 1.2-13.4) were associated with contracting MPVX infection.

**Conclusion:** Recent global outbreaks, the rising incidence in young adults and endemic zones might result from smallpox vaccine cessation. Increased risk with sick animal exposure or sleeping on the floor suggests high infectivity from animal excretions. Increasing awareness, strict surveillance, and contact tracing can contain global outbreaks. Ring vaccination approach to exposed people can also be a strategy. Future studies should investigate to determine measures for rapid laboratory diagnosis, maintaining lab safety, and also transmissibility.

## Introduction

As the world surpasses the shock of COVID-19 pandemic and learns lessons from the global health failures to control the global outbreak, new infectious diseases threats emerge such as human monkeypox (MPXV) virus infection. [1] Since May 7, 2022, the global health authorities were surprised to experience the unprecedented and unexpected MPVX outbreaks across Europe, the US, and Australia, which have continuously been reported in 12 WHO member states across three WHO regions. [1] MPXV infection, an emerging zoonotic disease caused by an orthopoxvirus, results in a clinically less severe smallpox-like disease. With the smallpox eradication in 1980 with subsequent cessation its vaccination, MPXV has emerged as the most concerning orthopoxvirus for public health [2]. In 1970, MPXV was first identified in a 9-month-old boy in the Democratic Republic of the Congo (DRC), where smallpox had been eliminated in 1968. Before April 2022, MPXV cases were endemic and seldom reported outside African regions where it primarily occurred in central and west Africa, often in proximity to tropical rainforests [2,3,4]. Epidemiologic investigations could not establish a travel link of reportedly rising cases with endemic areas. Animal hosts of this virus include a range of rodents and non-human primates and transmit to humans with an incubation period of 5 to 21 days. [1]

MPVX is an enveloped double-stranded DNA virus of the Orthopoxvirus genus of the Poxviridae family. [4] It has two distinct genetic clades, namely: the central African (Congo Basin) clade and the west African clade [4]. The Congo Basin clade has historically caused more severe disease and was thought to be more transmissible. The geographical division between the two clades has so far been in Cameroon, the only country where both virus clades have been found [4,5]. The true burden of MPVX cases have been unknown [6]. Since 2017, Nigeria has experienced a large outbreak with over 500 suspected and 200 confirmed cases with a 3% case fatality ratio. [7]

In 2003, the US experienced the first MPVX outbreak of 70 cases beyond Africa, which was linked to animal contact with infected pet prairie dogs [8]. These pets had been housed with Gambian pouched rats and dormice that had been imported into the country from Ghana [8]. Monkeypox has also been reported in travelers from Nigeria to Israel (2018), to United Kingdom (2018, 2019, 2021, 2022), to Singapore (2019), and to the US (2021). In May 2022, multiple cases of monkeypox were identified in several non-endemic countries as mentioned earlier [9]. MPVX infection has become a disease of global public health importance since it is globally spreading in an epidemic proportion.

The recent apparent increase in human MPVX cases across a wide geographic area, the potential for further spread, and the lack of reliable surveillance have heightened the concern for this emerging zoonosis. In November 2017, a collaborative consultation of the World Health Organization (WHO) and CDC with global researchers, global health partners, ministries of health, and orthopoxvirus experts had reviewed and discussed recently detected human MPVX infections in African countries and also identifed the surveillance and other improvement measures [10]. Endemic human monkeypox has been reported from more countries in the past decade than during the previous 40 years. Since 2016, confirmed cases of monkeypox have occurred in Central African Republic, Democratic Republic of the Congo, Liberia, Nigeria, Republic of the Congo, and Sierra Leone and in captive chimpanzees in Cameroon [11]. Many countries with endemic monkeypox lack recent experience and specific knowledge about the disease to detect cases, treat patients, and prevent further spread of the virus. Specific improvements in surveillance capacity, laboratory diagnostics, and infection control measures are needed to mount an efficient response [12]. The recent historic global outbreaks of MPXV following the shocking COVID-19 pandemic have raced scientists to contain it by investigating the change in epidemiology.

Therefore, we aimed to systematically review the current literature on evolutionary epidemiology of emerging human monkeypox virus infection and emphasize on the number of suspected, confirmed, and fatal cases as well as risk factors for contracting monkeypox infection.

## Methodology

### Search Strategy

We performed a systematic review of peer-reviewed literature in accordance with international standards for conducting and reporting systematic reviews including the Preferred Reporting Items for Systematic Reviews and Meta-Analyses (PRISMA) guideline. Searches were performed on three electronic databases including MEDLINE (accessed using PubMed), Online Willey Library, and Science Direct with no language restrictions. For monkeypox related studies, we included articles from 2018 to 2022 for better comparison.

The search string used the following keywords in all three databases “Orthopox virus” OR “Monkeypox virus” OR “Human Infection” and Transmission of virus to search relevant articles. We aimed to explore how monkeypox epidemiology is evolving including the data related to the number of suspected, confirmed, and fatal cases. We also sought to explore the risk factors for contracting monkeypox infection. We assured that all the data have information such as transmission, pathology and evolution and other features of this virus. Two independent researchers screened the titles and abstracts of the retrieved studies in electronic search and relevant studies were evaluated in full text for eligibility.

### Inclusion criteria

Peer-reviewed articles with complete demographic information and complete medical records of patients who were suffering from monkey pox were included.

### Exclusion Criteria

Non-English studies, letter to editors, posters, cases studies with incomplete information, non-human studies, modelling studies, case reports and articles with unavailable full text were excluded from the review.

### Data Extraction

All citations were imported into a bibliographic database and duplicates were removed. Title, abstract and then full text of all articles were screened for eligibility. All the studies which had directly included the MPXV transmission, reported outbreaks as well as sign and symptoms of MPXV were included. The following information was extracted from each study: a) details of the study (study setting, year of publication and study design), b) study population, sample size (male/female) and age in years, c) primary intervention (s) and control group and d) transmission of MPXV. The evaluation of selected data was further done into two phases, first, we selected the data based on abstract and title. Secondly, the full text of the articles was examined and included if they satisfied the eligibility criteria for inclusion.

## Results

The initial search strategy yielded a total of 762 articles, 164 of which were removed due to duplicated articles, and the remaining 598 articles were screened for full-text eligibility. On further screening, we omitted 502 articles with poor information. The remaining 96 articles were further screened for inclusion criteria and 25 articles that fulfilled the inclusion criteria and had adequate data were included. The PRISMA flowchart of the selection process for the systematic review is in shown in Figure 1.

**Fig 1:**
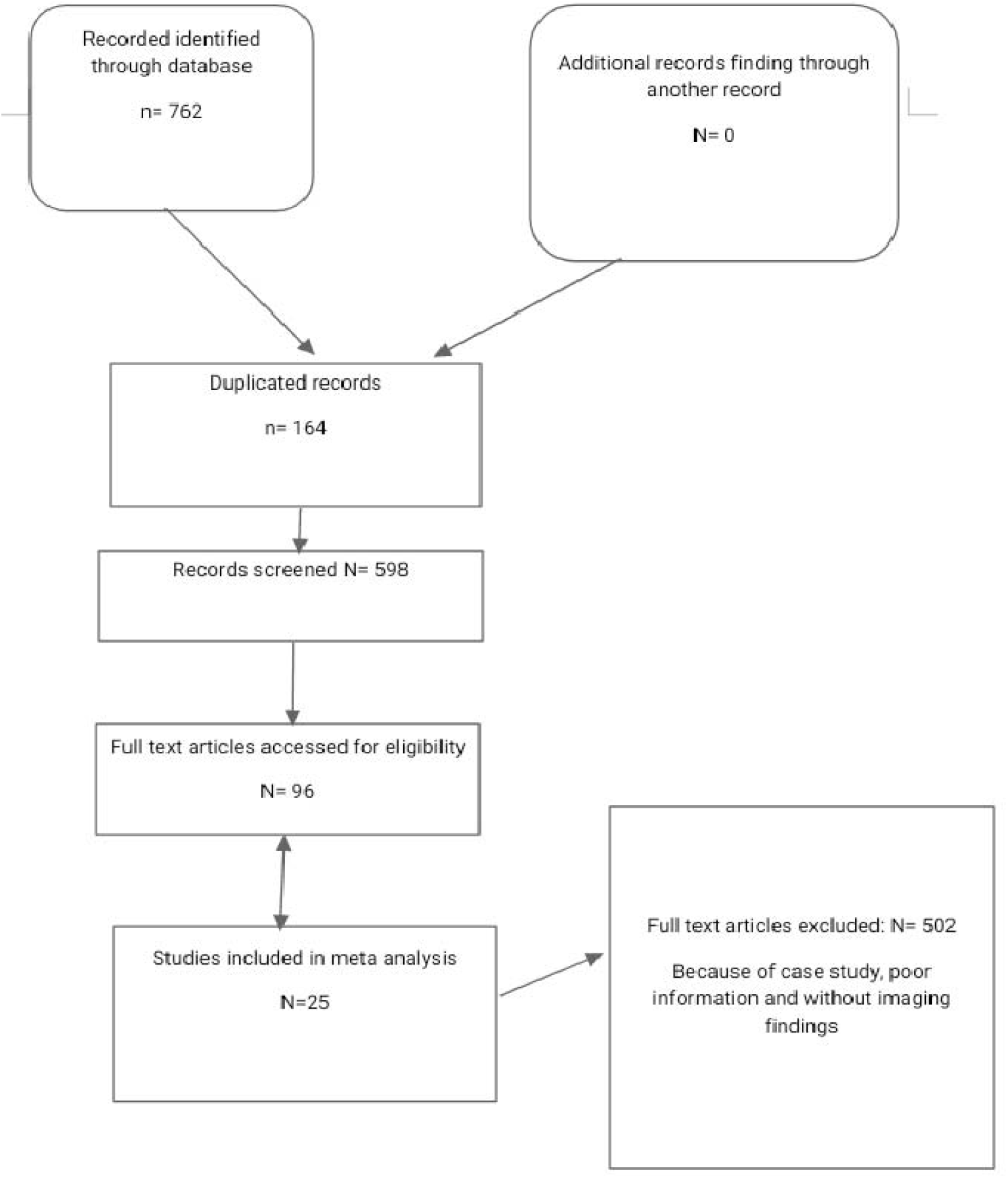
Inclusion Criteria of selected studies according to PRISMA follow up.

### Number of reports by country

Monkeypox data from the DRC accounted for more than one-third of the eligible articles [2,4, 8-11, 17-18, 21-22]. The remaining articles had monkeypox data from the CAR [4, 12, 24-25], Gabon [5, 7], Cameroon [4, 6, 23], ROC [13, 16, 19, 29], Sierra Leone [3, 20], Cote d’Ivoire [3-4], Nigeria [3, 30], and Liberia [3, 31]. (Note: two articles [3-4] described data for more than one country, therefore the total number of articles per country exceeds 25.

### Number of cases by country

We identified 25 peer-reviewed articles [1-12, 15-24, 28-30] with data on the number of confirmed cases, number of deaths and case fatality rate (CFR) from human MPXV infection. Since the beginning of 2022, the continent has reported 4,667 cases (559 confirmed; 4,108 suspected) and 127 deaths (CFR: 2.8%) of monkeypox from eight endemic MS: Benin (3 suspected; 3 confirmed; 0 deaths), Cameroon (29; 7; 2), CAR (17; 8; 2), Congo (14; 5; 3), DRC (2,775; 163; 110), Ghana (535; 84; 4), Liberia (31; 2; 0), Nigeria (704; 277; 6) and four non-endemic MS: Egypt (0; 1; 0), Morocco (0; 3; 0), South Africa (0; 5; 0) and Sudan (0; 1; 0). This week, a total of 423 new cases (39 new confirmed; 384 new suspected) and three new deaths of monkeypox were reported from Benin, Congo, Ghana and Liberia.

### Outbreak investigation of monkeypox virus infection between 2018-2022

A systemic review and meta-analysis (Nikola et al., 2018) that aimed to study human MPXV outbreaks in Congo showed the urgent need to increase surveillance and preparedness capacity to prevent and control the outbreaks. Another review (Kara et al., 2018) which investigated the emerging trends of MPXV infections in South Africa suggest the need for a one health approach for disease detection as well as wildlife surveillance and investigations into the animal reservoir/reservoirs. [Table 2]

**Table 1.** Suspected, confirmed and fatal monkeypox cases by country and year. [Each box denotes number of probable cases, number of confirmed cases (n), and number of deaths, and/or CFR (%). Where suspect cases were tested and found positive for another disease, these were subtracted from the suspect case total. CFR calculated as number of deaths out of total suspect cases. Grey denotes a period without reported outbreaks. Green denotes locations affected by CB clade. Orange denotes areas affected by WA clade.]

| Country | DRC (Zaire 1971-97)* | South Sudan (Sudan pre-2011) | Gabon | Cameroon | CAR | ROC | Sierra Leone | Cote d'Ivoire | Nigeria | Liberia |
| --- | --- | --- | --- | --- | --- | --- | --- | --- | --- | --- |
| 1970-1974 | 1970: 1 (1)<br>100% [2]<br>1971: 0<br>1972: 5<br>1973: 3<br>1974: 1 |  |  |  |  |  | 1970-71:<br>1 (1) 0%<br>[3] | 10/1971:<br>1 (U)<br>0% [3] | 1971:<br>2 (1)<br>0% [3] | 1970-71:<br>4 (1)<br>0% [3] |
| 1975-1979 | 1975: 3<br>1976: 5<br>1977: 6<br>1978: 12<br>1979: 8 |  |  | 1979: 2 (U)<br>U% [4] |  |  |  |  | 1978:<br>1 (1)<br>0% [3] |  |
| 1980-1984 | 1980: 4<br>1981: 7<br>1982: 40<br>1983: 84<br>1984: 86 |  | 1987:<br>Lambarene:<br>1 (1)<br>100% □ □<br>[5] |  | 1984: 6 (U)<br>U% [4] |  |  | 1981: 1<br>(U) U%<br>[4] |  |  |
| 1985-1989 | 1985: 62<br>1986: 59 [4]<br>1981-86: 338<br>(U) 9.8% [4] |  |  | 1989: 1 (1)<br>0% [6] |  |  |  |  |  |  |
| 1990-1994 |  |  | 01/1991-06/1991: Region between Lamberene and N'Djole: 9 (5) 0% [7] |  |  |  |  |  |  |  |
| 1995-1999 | 02/1996-02/1997: Katako-Kombe HZ: 92 (12) 3.3%<sup>††</sup> [8]<br>02/1996 – 10/1997: Kasai Oriental: 511 (U) (1.5%)<sup>§§</sup> [9]<br>03/1997 - 05/1997: Katako-Kombe n=112, Lodja Nord n=58, Sud HZ n=11: 170 (U) 0% (Weekly Epidemiological Record) [10] |  |  |  |  |  |  |  |  |  |
| 2000** -2004 | 02/2001-08/2001: Equateur Province: 23 (9) 21.7% [11]<br>2001: 388 (U) n=13 (3.4%)<br>2002: 881 (1.6%)<br>2003: 755 (2.1%)<br>2004: 1024 (2.8%) |  |  |  | 14/08/2001: Pimu CAR/DRC border: 8 cases in a family (U) 25% [12]<sup>††</sup> | 15/04/03 – 23/06/2003: Likouala department: 12 (3) 8.3%<sup>†</sup> [13] |  |  |  |  |
| 2005-2009 | 2005: 1708 (1.5%)<br>2006: 783 (2.6%)<br>2005-7: Sankuru District: 1407 (703) U% [13]<br>2007: 970 (1.1%)<br>2008: 1599 (4.2%)<br>2009: 1919 (1.4%) | 10/2005: Unity State: 37 (10) 0% [14] |  |  |  | 2007: Likouala department: 62 - 150+ (U) U%<sup>‡</sup> (IRIN, Reliefweb) [15] |  |  |  |  |
| 2010-2014 | 2010: 2322 (1.1%)<br>2011: 2208 (0.7%)<br>2012: 2629 (1.3%)<br>2013: 2460 (1.5%)†† (IDSR data) [16]<br>2013: Bokungu HZ: 99 (50) 10 (10.1%) [17]¶ |  |  |  | 06/2010: Deep forest, Southern CAR, 480km from DRC border: 2 (2) 0%§ [11]<br>30/04/2012: Batangafo: 2 (U) U% [11] | 04/10-11/2010: Likouala department: 11 (2) 9.1% [18] | 2014: Bo District: 1 (U) U% [19] |  |  |  |
| 2015-2018 | 1/1/2016 - 1/3/2016: Ateki HZ: 155 (7) 11 (7.1%) [21]<br>01/01/2018 - 08/07/2018: IDSR: 2995 (U) 1.2% [22] |  |  | 30/04/2018: Njikwa Health District: 6 (1) 0% [23] | 30/01/2015: Bria: 3 (U) n=1 33%<br>04/12/2015 - 02/2016: Bakouma and Bangassou subprefectures, Mbomou province: 10 (3) 20% [24]<br>04/12/2015-28/09/2016: Additional 3 cases reported□ (IFRC) [25]<br>04/09/16-07/10/2016: Haute-Kotto health district: 26 (4) | 18/01/17 – 15/10/17: Likouala department: 88 (7) 6.8% [29] | 14/03/2017: Pujehun district: 1 (1) 0% [20] |  | 2017-18: 24 States: 228 (89) 2.6% [30] | 12/2016 1 (U) U% [31] |

**Table 02.** Summary of study findings on monkeypox virus infection outbreaks between 2018 to 2022

| Author;<br>Published Year | Objective | Study design | Study population | Outcomes |
| --- | --- | --- | --- | --- |
| Nikola et al., 2018 | Trace all reported human monkeypox outbreaks and relevant epidemiological information | Systematic review analysis | Congo | urgent need to focus on building surveillance capacities which will provide valuable information for designing appropriate prevention, preparedness and response activities. |
| Kara et al., 2018 | MPX virus as an emerging trend in humans | Review analysis | South Africa | As with all zoonotic diseases, a comprehensive One Health<sup>8</sup> approach is necessary for disease detection and response, including wildlife surveillance and investigations into the animal reservoir/reservoirs, which require dedicated resources. |
| James et al., 2021 | Zoonotic orthopoxvirus outbreaks | Review analysis | Middle East and India, buffalopox in India, vaccinia in South America, and novel emerging orthopoxvirus infections in the United States, Europe, Asia, and South America | outbreaks have occurred repeatedly worldwide |
| Hala et al., 2022 | Monkeypox virus: A spreading threat for Pakistan | Review analysis | Pakistan | despite public-health experts declaring Monkeypox a 'containable disease' (unlike COVID-19), the Pakistani government and health ministries must take this matter seriously and adequately prepare for a potential outbreak. |

### Mode of transmission and risk factors

In DRC, sleeping in the same floor was significantly (odds ratio [OR] 6.1, 95% confidence interval [CI] 1.2-31.6) associated with contracting MPVX while eating or processing animal foods was not a significant risk factor. In United States, cleaning cages and bedding a sick animal (OR 5.3, 95% CI 1.4-20.7), or touched an infected animal (OR 4.0, 95% CI 1.2-13.4), daily direct or indirect sick animal exposure (OR 4.0, 95% CI 1.2-13.4) were associated with contracting MPVX infection. [Table 3]

**Table 03.** Summary of study findings on risk factors for primary introduction of monkeypox.

| Country | Risk Factor | OR | aOR for vaccination status (95% CI) | P Value | Number of cases, n (Number of controls, n) |
| --- | --- | --- | --- | --- | --- |
| DRC [2]* | Vaccinated | 0.1 (0.03, 0.6) | NA | ND | 252 (653) |
|  | Exposure to Gambian rats | ND | 2.6 (1.6, 4.1) | ND |  |
|  | Large terrestrial rodents | ND | 1.8 (1.1, 3.0) | ND |  |
|  | Prosimians | ND | 1.9 (1.2, 2.8) | ND |  |
|  | Non-human primates | ND | 2.7 (1.4, 4.9) | ND |  |
| DRC [3] | Live in house with a door | 0.07 (0.01, 0.6) | ND | 0.01 | 15 (50) |
|  | Prepared wild animal for consumption | 0.2 (0.09, 0.1) | ND | 0.04 |  |
|  | Ate duiker | 0.15 (0.03, 0.7) | ND | 0.01 |  |
|  | Sleep on floor | 6.1 (1.2, 31.6) | ND | 0.03 |  |
| USA [4] | Vaccinated | 0.3 (0.1, 0.9) | NA | ND | 30 (35)<sup>†</sup> |
|  | Touched an infected animal | 3.8 (1.2–11.7) | 4.0 (1.2, 13.4) | ND |  |
|  | Cleaned the cage or touched the bedding of an infected animal | 5.3 (1.5, 18.9) | 5.3 (1.4, 20.7) | ND |  |
|  | Received scratch from infected animal | 5.6 (1.1, 28.6) | 3.9 (0.7, 21.1) | ND |  |
|  | Daily indirect or direct exposure to an ill animal was significantly associated with MPX developing | 3.8 (1.2, 11.7) | 4.0 (1.2, 13.4) | ND |  |
|  | Having come within 6 feet but not touched by an infected animal | 2.0 (0.6, 6.2) | 2.0 (0.6, 6.5) | ND |  |

NA = Not applicable ND = Not Described \*Conference abstract, no methodology provided <sup>†</sup>Three controls were found to have elevated levels of IgM to orthopoxvirus.

## DISCUSSION

This systematic review provides a comprehensive review of the epidemiologic evolution of MPXV infection since its first human detection in 1970s. We found a greater than 14-fold increase in confirmed MPXV cases over the past 5 decades, from 55 cases in the 1970s to 793 cases in the 1990s, with DRC being the most affected country. As a result of the recent outbreak, the number of confirmed cases in Nigeria has escalated from 3 cases in the 1970s to 341 cases in 2015–2018. There is a growing concern regarding the geographical spread and re-emergence of MPXV. In last five decades, the outbreaks have been reported in 10 African countries. Concerningly, the re-emergence of MPXV occurred in Nigeria, Liberia, and Sierra Leone after 40 years and in the CAR after 30 years. We report a rapid rise of MPVX cases in a highly endemic DRC after 1970s while spreading to other countries. Cessation of smallpox vaccination might be one of the factors responsible for the findings.

Monkeypox typically transmits to humans from wild animals, such as rodents and primates (animals-to-humans), or in some cases, from other infected individuals (humans-to-humans) [4, 36]. Nine studies showed the source of infection was sexual contact, especially with male partners. Six studies mentioned the cause of infection was contact with an individual with monkeypox symptoms. Two studies considered cases due to acquired nosocomial infection. Ingestion of barbecued bushmeat was the source of infection in three studies and rodent carcasses were the source of infection in the other two studies [40]. The opportunistic infection predominantly spreads through contact, such as hunting or consuming disease-ridden animals [4, 36]. It can also spread through large aerosol droplets or contact with body fluids (saliva and blood), infected skin lesions, or contaminated materials, such as the clothing of an infected individual [4, 37]. Following an incubation period of 5–21 days, the virus clinically manifests as fever, headache, myalgia, lymphadenopathy, and fatigue, similar to the presentation of the smallpox infection [4,37]. Individuals infected with a more barbaric form of the disease may also develop a characteristic *‘Monkeypox rash’*, starting from their face and hands and later spreading to other body parts [5,38]. The lesions which typically start as macules progress into papules, followed by fluid-filled vesicles, pus-filled pustules, and subsequently, scabs that crust and eventually fall off [5,38]. The disease is often self-limiting, with the symptoms eventually resolving within 2–4 weeks. As supported by historical data, the smallpox vaccine (Imvanex) is 85% effective against Monkeypox. Similarly, antivirals, including Tecovirimat, Cidofovir, and Vaccinia Immune Globulin Intravenous (VIGIV) approved for smallpox infection, can also be used for Monkeypox treatment [36,37].

Due to the atypical disease presentation and transmission patterns, the recent MPXV surge is “rare and unusual” compared to earlier outbreaks. [36,37] As reported by health experts, most of the Monkeypox cases recorded in the past month in the Western Hemisphere have no known epidemiological links to endemic countries [37]. Additionally, most of the cases were documented among young men engaging in homosexual or bisexual activities, commonly known as “men who have sex with men” (MSM) [37]. Most of these cases presented with genital or peri-genital lesions, indicating a new route of disease spread, sexual contact. Although a definite cause for the sudden upsurge in cases is yet to be determined, health experts worldwide have suggested several theories which are currently under investigation. Till 2022 genomic sequences of ten MPXV strains, associated with the recent countrywide outbreak, have been determined. While West African Clade is mostly implicated in the recent viral surge, data were insufficient to determine which mutation contributed to increased transmissibility [38,39]. We found that in DRC, sleeping on the floor was significantly associated with contracting MPXV while eating or processing animal foods was not a significant risk factor. In the United States, cleaning cages, bedding sick animals, or touching infected animals, daily sick animal exposure was associated with contracting MPVX infection.

Health officials stopped recommending smallpox inoculation following the eradication of smallpox in the late nineties [36,37]. Given that smallpox and Monkeypox viruses belong to the same genus (Orthopoxvirus), the cessation of the smallpox vaccine resulted in populations developing a weakened immunity to MPXV [37]. The waning immunity against the virus over several years served as a nidus for infection, resulting in the latest re-surge. almost five decades ago, MPXV was considered a “disease of young children”, predominately affecting children aged 4–5 years [37]. By the 2000s, this number had increased five times, with ages 20–25 years old being most vulnerable to infection [38,39]. This is in line with the current data, which suggests that most of the recently reported MPXV cases have been in young men. It is highly likely that most of these individuals were not administered the smallpox vaccine either because they were too young or were born after the termination of routine vaccination.

The resumption of everyday life following the Coronavirus-2019 (COVID-19) pandemic can also be attributed as a factor favoring MPXV spread. With the lifting of travel restrictions coupled with the easing of social distancing measures, individuals were more likely to have been in close physical contact, thereby encouraging viral transmission. Moreover, the decline in the number of dendritic cells, and consequently the wearying immune system response post-COVID-19 infection, results in recovered individuals being more vulnerable to secondary infections; the Monkeypox re-surge might be just another testament to this fact.

## Conclusion

Recent global outbreaks, the rising incidence in young adults and endemic zones might result from smallpox vaccine cessation. Increased risk with sick animal exposure or sleeping on the floor suggests high infectivity from animal excretions. Increasing awareness, strict surveillance, and contact tracing can contain global outbreaks. Ring vaccination approach to exposed people can also be a strategy. Future studies should investigate to determine measures for rapid laboratory diagnosis, maintaining lab safety, and also transmissibility. In addition, given that diagnosing MPXV infection based on their symptoms is not a reliable approach, it is imperative that the WHO urgently procures the required testing kits, primers, and reagents to combat a possible viral epidemic.

## Recommendations

Raising awareness among healthcare providers and the general population, implementing strict surveillance measures and timely contact tracing (in case of a viral outbreak) are the best preventative measures to effectively prevent or manage a potential Monkeypox outbreak in the country. Per expert-issued guidelines, mass smallpox vaccination campaigns are not a requirement. Instead, a ring vaccination approach needs to be utilized, that is, to only vaccinate people in close contact with infected individuals. This way the challenges presented due to the limited global supply of smallpox vaccine and the prevalent vaccination hesitancy in the world can be overcome, albeit to some extent.

## Data Availability

All data produced in the present work are contained in the manuscript

## Acknowledgments

Not applicable.

## Declarations

Ethics approval and consent to participate not applicable.

## Author Contributions

Sunder Sham: Study design, collecting data, data analysis, and manuscript writing.

FNU Sapna, Sanjay Kumar, Anjali, Ahmed Bandari, Reham Al-Refai, Mohammed Abdelwahed, Manal Mohammad Baloch, Soumya Jaladi, FNU Kiran and Hansini Laharwani, and data collection and analysis

Vivek Podder: review and editing

All the authors have read and approved the final manuscript.

